# Intronic *FGF14* GAA repeat expansions are a common cause of downbeat nystagmus syndromes: frequency, phenotypic profile, and 4-aminopyridine treatment response

**DOI:** 10.1101/2023.07.30.23293380

**Authors:** David Pellerin, Felix Heindl, Carlo Wilke, Matt C. Danzi, Andreas Traschütz, Catherine Ashton, Marie-Josée Dicaire, Alexanne Cuillerier, Giulia Del Gobbo, Kym M. Boycott, Jens Claassen, Dan Rujescu, Annette M. Hartmann, Stephan Zuchner, Bernard Brais, Michael Strupp, Matthis Synofzik

## Abstract

The cause of downbeat nystagmus (DBN) remains unknown in approximately 30% of patients (idiopathic DBN). Here, we hypothesized that: (i) *FGF14* (GAA)_≥250_ repeat expansions represent a frequent genetic cause of idiopathic DBN syndromes, (ii) are treatable with 4-aminopyridine (4-AP), and (iii) *FGF14* (GAA)_200-249_ alleles are potentially pathogenic.

We conducted a multi-modal cohort study of 170 patients with idiopathic DBN that comprised: in-depth ocular motor, neurological, and disease evolution phenotyping; assessment of 4-AP treatment response, including re-analysis of placebo-controlled video-oculography treatment response data from a previous randomized double-blind 4-AP trial; and genotyping of the *FGF14* repeat.

Frequency of *FGF14* (GAA)_≥250_ expansions was 48% (82/170) in the entire idiopathic DBN cohort. Additional cerebellar ocular motor signs were observed in 100% (82/82), cerebellar ataxia in 43% (35/82), and extracerebellar features in 21% (17/82) of (GAA)_≥250_-*FGF14* patients. Alleles of 200 to 249 GAA repeats were enriched in patients with DBN (12%; 20/170) compared to controls (0.87%; 19/2,191; OR, 15.20; 95% CI, 7.52-30.80; *p*=9.876e-14). The phenotype of (GAA)_200-249_-*FGF14* patients closely mirrored that of (GAA)_≥250_-*FGF14* patients. (GAA)_≥250_-*FGF14* and (GAA)_200-249_-*FGF14* patients had a significantly greater clinician-reported (80% vs 31%; *p*=0.0011) and self-reported (59% vs 11%; *p*=0.0003) response rate to 4-AP treatment compared to (GAA)_<200_-*FGF14* patients. This included a treatment response with high relevance to everyday living, as exemplified by an improvement of 2 FARS stages in some cases. Placebo-controlled video-oculography data of four (GAA)_≥250_-*FGF14* patients previously enrolled in a 4-AP randomized double-blind trial showed a significant decrease in slow phase velocity of DBN with 4-AP, but not placebo.

This study shows that *FGF14* GAA repeat expansions are a highly frequent genetic cause of DBN syndromes, especially when associated with additional cerebellar features. Moreover, they genetically stratify a subgroup of patients with DBN that appear to be highly responsive to 4-AP, thus paving the way for a “theranostics” approach in DBN syndromes.

## Introduction

Downbeat nystagmus (DBN) is the most common form of acquired persisting nystagmus and is characterized by a pathological slow upward drift of the eyes followed by a corrective downward saccade.^1,2^ In most cases it is caused by a hypofunction of the cerebellar flocculus/paraflocculus or their projections, which leads to disinhibition of the superior vestibular nuclei neurons, resulting in spontaneous slow upward eye drift.^3-6^ DBN is a common feature of the allelic disorders spinocerebellar ataxia 6 and episodic ataxia type 2, while it is infrequently observed in other forms of spinocerebellar ataxia.^7-10^ Despite the numerous known genetic and acquired etiologies,^1,2^ the cause of DBN remains unknown in approximately 30% of cases (idiopathic DBN).^2^

The recent identification of an association between a variation (rs72665334) in intron 1 of the fibroblast growth factor 14 (*FGF14*) gene and idiopathic DBN in a genome-wide association study (GWAS)^11^ and of a dominantly inherited (GAA)_≥250_ repeat expansion in intron 1 of *FGF14* as the cause of spinocerebellar ataxia 27B (SCA27B) / GAA-*FGF14* ataxia,^12,13^ a late-onset slowly progressive cerebellar syndrome that is frequently associated with DBN,^12,14^ provided evidence for a potential link between *FGF14* and DBN.^12,14,15^ Furthermore, 4-aminopyridine (4-AP), a proven effective symptomatic treatment for DBN,^16,17^ has shown promising benefits to reduce the frequency and severity of ataxic symptoms in previous small series of patients with GAA-*FGF14* ataxia.^12,14,18^

Based on these findings, we hypothesized that *FGF14* (GAA)_≥250_ repeat expansions are a frequent monogenic cause of idiopathic DBN syndromes, which are treatable with 4-AP, and are in disequilibrium with the genome-wide significant rs72665334 variant previously identified by GWAS. Moreover, we systematically studied the phenotype and 4-AP treatment response of patients with DBN carrying an *FGF14* (GAA)_200-249_ allele, providing preliminary evidence that these so-called “intermediate” alleles might be pathogenic.

## Methods

### Patient Enrollment

We enrolled a consecutive series of 219 index patients with suspected DBN referred to the Department of Neurology or the German Center for Vertigo and Balance Disorders at the LMU Hospital in Munich, Germany, between 2012 and 2020. Etiologic evaluation of DBN was performed through a detailed medical history, assessment of drug abuse, comprehensive neurological examination, laboratory tests, and brain imaging by MRI or CT scan. Genetic screening for episodic ataxias and spinocerebellar ataxias was performed in select patients with a suggestive phenotype when deemed appropriate by the clinician. Patients were excluded from the study if: (i) no DNA was available for genetic screening (*n*=2), (ii) DBN was not objectified on neuro-ophthalmological examination after review of the medical records (*n*=11), or (iii) a competing etiology for DBN was identified (*n*=36). The final study cohort comprised 170 patients with a diagnosis of idiopathic DBN (**Fig. 1**). Part of this patient cohort (*n*=80) has previously been reported in the idiopathic DBN GWAS.^11^ All but three patients (one [GAA] -*FGF14* and two [GAA] -*FGF14*) were of self-reported European descent. The three non-European patients were of Turkish descent. This study was approved by the ethics committees of the LMU Munich, the Montreal Neurological Hospital-Institute, and Clinical Trials Ontario and was carried out in accordance with the Declaration of Helsinki. We obtained written informed consent from all the participants in this study.

**Figure 1:**
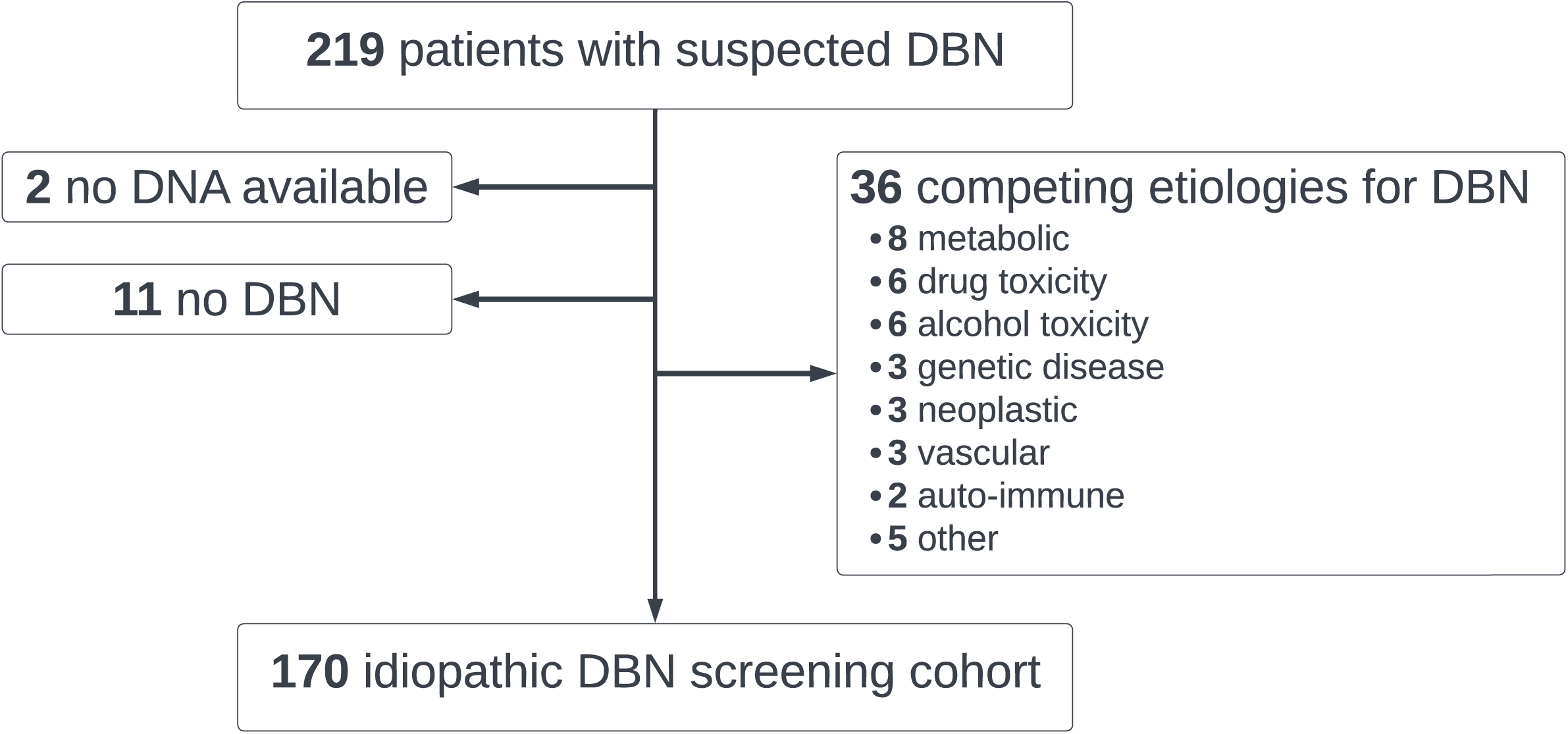
Study flowchart of the recruitment of patients with idiopathic DBN. DBN: downbeat nystagmus.

### Deep Phenotyping

All patients underwent at least one comprehensive neurological and ocular motor examination. Deep phenotyping was performed by systematically reassessing all medical records using a standardized data sheet while being blind to the GAA-*FGF14* genotype. Functional impairment was assessed in terms of the need for mobility aids and by means of the Friedreich Ataxia Rating Scale (FARS) functional stage (0=normal; 1=minimal signs on examination; 2=minimal disability; 3=mild disability; 4=moderate disability, requires a walker; 5=severe disability, confined but can navigate a wheelchair; 6=total disability).^19^ Results of routine brain MRI and nerve conduction studies were available for review in 70% (119/170) and 18% (31/170) of patients, respectively. Ancillary assessment of vestibular function was obtained in 94% (159/170) of patients using bithermal caloric stimulation (*n*=147), video head impulse test (vHIT; *n*=100), and rotatory chair test (*n*=1). Posturography (Kistler platform, Kistler Instrumente AG) was obtained in 59% (100/170) of patients and results were analyzed with an artificial neural network.^20^

Patients were stratified in one of four phenotypic clusters based on the presence of additional cerebellar and/or extracerebellar signs: pure DBN, DBN plus additional isolated cerebellar ocular motor signs (DBN+COM), DBN plus cerebellar ataxia (DBN+CA), and DBN plus cerebellar and extracerebellar features (DBN+EC), which included bilateral vestibulopathy (BVP) and/or polyneuropathy. Along with the cerebellar system, the vestibular and sensory systems appear to be preferentially impaired in GAA-*FGF14* ataxia.^14,15^ Additional cerebellar ocular motor signs were defined by the presence of at least one of: saccadic pursuit, dysmetric saccades, gaze-evoked nystagmus, rebound nystagmus, or impaired visual fixation suppression of the vestibulo-ocular reflex (VOR). Cerebellar ataxia was defined by the presence of cerebellar dysarthria, dysdiadochokinesia, intention tremor, or, when available, evidence of cerebellar involvement on posturography. BVP was diagnosed as per the consensus criteria of the Bárány Society requiring the documentation of bilaterally reduced or absent angular VOR function by caloric stimulation, vHIT, or rotatory chair.^21^ Polyneuropathy was diagnosed on nerve conduction studies (excluding focal entrapment neuropathies) or clinically defined by the combination of significantly decreased vibration sense at the ankles (≤3/8 on the Rydel-Seiffer scale) and decreased or absent ankle reflexes.^22^

### Assessment of Treatment Response to 4-aminopyridine

Assessment of treatment response to 4-AP was performed by two independent approaches. (1) *Open-label real-world treatment response data*. Information on clinician-reported and patient-reported response to 4-AP treatment (fampridine 10mg twice a day) was collected (available for 83 patients). As all patients from this study who were treated with 4-AP received the drug before the discovery of GAA-*FGF14* ataxia, the assessment of treatment response was naturally blind to the underlying GAA-*FGF14* genotype. Patients were routinely evaluated 1 to 3 months after initiating treatment with 4-AP. Patient-reported responses were assessed based on their global impressions of the impact of treatment on their neurological functioning. Clinician-reported responses were evaluated based on the clinician’s global impression from neurological and neuro-ophthalmological examinations. (2) *Double-blind placebo-controlled clinical trial data*. Four patients with DBN had been part of an earlier placebo-controlled randomized double-blind trial assessing the efficacy of 4-AP in a cohort of 27 patients with DBN.^16^ All four patients, now known via the current study to carry an *FGF14* (GAA)_≥250_ expansion, were genetically undiagnosed at the time of the original trial.^16^ Due to unavailability of DNA, the genetic status of the remaining 23 patients who took part in the 4-AP trial remains unknown. As part of the trial, patients had received 5 mg of 4-AP (or placebo) four times a day for 3 days and 10 mg of 4-AP (or placebo) four times a day for 4 days. Randomization resulted in all four patients receiving active treatment first, then placebo, separated by a 1-week washout. Assessments were done before the first, 60 min after the first, and 60 min after the last drug administration, measuring the slow phase velocity (SPV) of DBN by video-oculography (degrees/second) as primary outcome.

### Genetic Screening for *FGF14* Repeat Expansions

The *FGF14* repeat locus was genotyped by capillary electrophoresis of fluorescent long-range PCR amplification products and bidirectional repeat-primed PCRs, as described previously.^23^ Expansions of at least 250 GAA repeat units were considered pathogenic.^12,13^ Alleles of 200 to 249 GAA repeat units were analyzed separately as their pathogenicity has recently been suggested.^24^

### Genotyping of the *FGF14* rs72665334 Variant

The rs72665334 C>T variant in intron 1 of *FGF14* (GRCh38, chr13:102,150,076) was found to be associated with idiopathic DBN in 106 patients in a recent GWAS.^11^ Genotyping data were previously generated using the HumanOmniExpress-24 array and data for the rs72665334 variant with call rate probability >70% were extracted for 73 patients enrolled in this study.

To assess whether the rs72665334 variant is in disequilibrium with the *FGF14* GAA repeat expansion at a population level, we also genotyped the rs72665334 variant by Sanger sequencing in an independent and ethnically distinct cohort of 37 French-Canadian index patients with GAA-*FGF14* ataxia and DBN. PCR reactions were performed in a 24µL volume using the Qiagen Taq DNA polymerase kit (catalog no. 201209, Qiagen) with 0.125mM dNTPs, 1µM of forward and reverse primers (forward primer: 5’-GCCCCTGTTCTAAAGCCTCT-3’; reverse primer: 5’-GATCGTCCAGCCACATCTCT-3’), and 240ng of genomic DNA. Sanger sequencing of PCR amplification products was performed using the ABI 3730*xl* Analyzer (Applied Biosystems).

### Statistical Analysis

We assessed differences between groups with the non-parametric Mann-Whitney U test for continuous variables and the Fisher’s exact test for categorical variables. Effect sizes are reported as odds ratio (OR) with 95% confidence intervals (CIs). The non-parametric Kruskal-Wallis test was used for comparisons between multiple groups. Correlations were calculated using the Pearson’s correlation coefficient or the Spearman’s rank correlation coefficient. The Kaplan-Meier method was used to analyze disease-free survival. Cross-sectional FARS stages measured off 4-AP were analyzed by linear regression as a function of disease duration, age, and the expansion size. To estimate the longitudinal progression of disability over the disease course, we used a linear mixed-effects model fitted by the restricted maximum likelihood method accounting for disease duration as fixed effect and with random intercepts and random slopes. We re-analyzed treatment effects of 4-AP and placebo by means of a mixed-effect analysis of repeated measures, with post-hoc comparison by uncorrected Fisher’s least significant difference. We analyzed the data in R (version 4.3) and GraphPad Prism 9.3. *P* value of <0.05 was considered significant, using the Benjamini-Hochberg method to correct for multiple comparisons. All analyses were two-sided.

### Data Availability

Individual deidentified patient data may be shared at the request of any qualified investigator upon reasonable request. No consent for open sharing has been obtained.

## Results

### Frequency of *FGF14* GAA Repeat Expansions in Downbeat Nystagmus Syndromes

A total of 170 index patients with a diagnosis of idiopathic DBN formed the final study cohort and were screened for *FGF14* GAA repeat expansions (**Figs. 1 and 2A**). We identified 82 patients (48%) who carried an *FGF14* (GAA)_≥250_ expansion (median size of expansion, 324 repeat units; range, 250-516), including one patient carrying biallelic GAA expansions of 280 and 313 repeat units. Alleles of 200 to 249 repeat units were identified in 12% of patients (20/170; median size of allele, 234 repeat units; range, 207-249), compared to 0.87% in 2,191 previously reported control individuals^25^ (19/2,191), suggesting an enrichment of this population of alleles in patients with DBN (OR, 15.20; 95% CI, 7.52-30.80; Fisher’s exact test, *p*=9.876e-14). Two patients carried a likely non-pathogenic^24,26^ (GAAGGA)_n_ expansion of 319 and 335 triplet repeat units equivalent, respectively.

**Figure 2:**
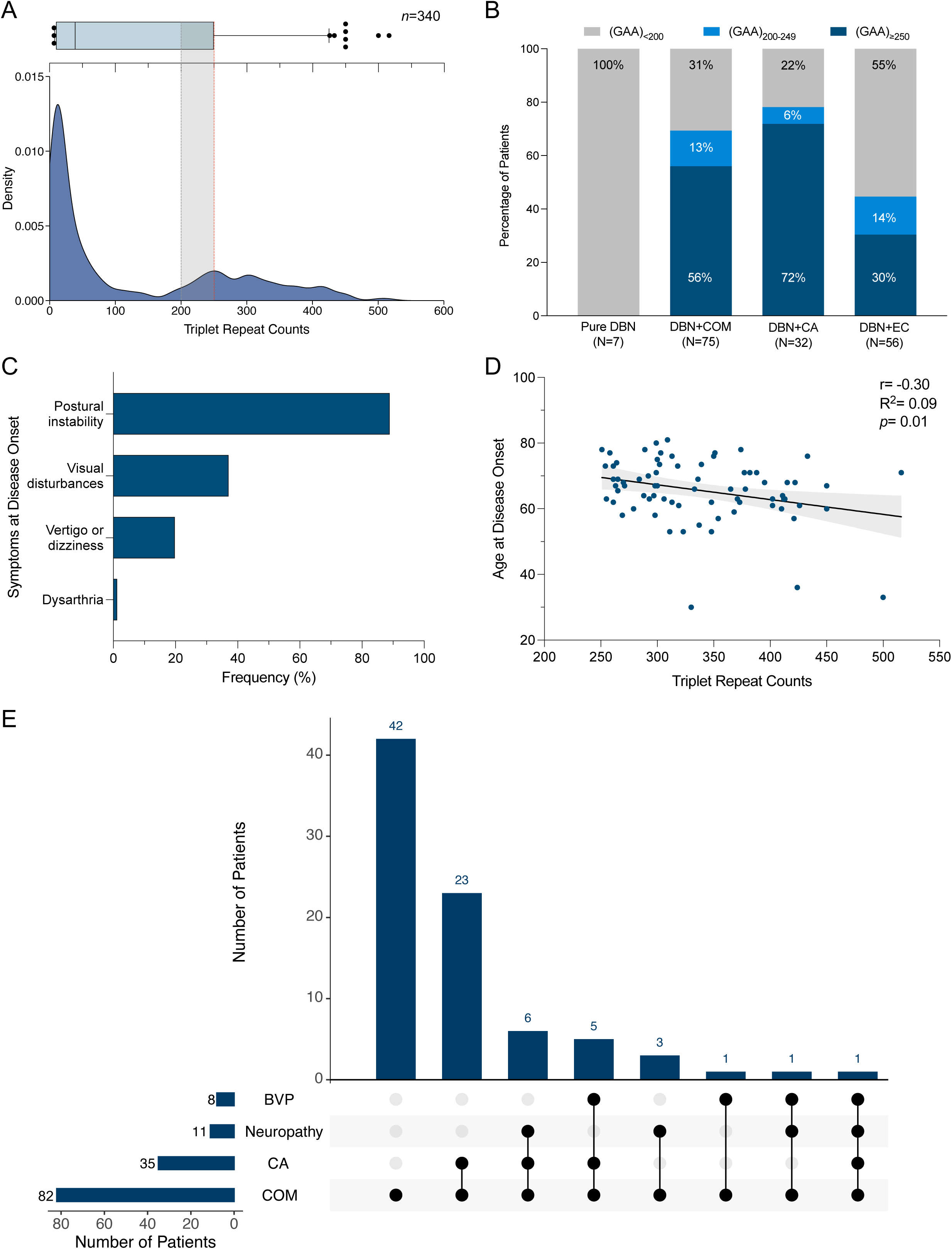
Frequency of *FGF14* GAA repeat expansions in DBN syndromes. (**A**) Allele distribution of the *FGF14* repeat locus in 170 patients with idiopathic DBN syndromes (340 chromosomes). The density plot shows allele size frequencies, with higher densities indicating greater frequencies. The box-and-whisker plot above the graph shows the allele distribution. The box indicates the 25^th^ percentile (first quartile), the median, and the 75^th^ percentile (third quartile), and the whiskers indicate the 2.5^th^ and 97.5^th^ percentiles. Outliers are represented by black dots. The dashed gray line and the shaded gray area indicate the so-called “intermediate” allele range of (GAA)_200-249_, and the dashed red line represents the pathogenic threshold of (GAA)_≥250_ repeat units. (**B**) Percentage of patients carrying an *FGF14* (GAA)_≥250_ expansion (dark blue), an *FGF14* (GAA)_200-249_ allele (light blue), and an *FGF14* (GAA)_<200_ allele (gray) in the subgroups with (1) pure DBN (0/7, 0/7, and 7/7 patients, respectively), (2) DBN plus additional isolated cerebellar ocular motor signs (DBN+COM) (42/75, 10/75, and 23/75 patients), (3) DBN plus cerebellar ataxia (DBN+CA) (23/32, 2/32, and 7/32 patients), and (4) DBN plus cerebellar ocular motor signs and/or ataxia and extracerebellar features (DBN+EC) (17/56, 8/56, and 31/56 patients). (**C**) Frequency of presenting symptoms in 81 patients with DBN carrying an *FGF14* (GAA)_≥250_ expansion. Data on presenting symptoms were missing for one patient. Patients may present with multiple symptoms at disease onset. Visual disturbances include diplopia, oscillopsia, and visual blurring. (**D**) Weak inverse correlation between size of the repeat expansion and age at disease onset in 74 patients carrying an *FGF14* (GAA)_≥250_ expansion for whom information on age at onset was available (Pearson’s r, -0.30; R^2^=0.09; *p*=0.01). The gray area displays the 95% confidence interval. (**E**) UpSet plot showing co-occurrence of cerebellar ocular motor signs (COM), cerebellar ataxia (CA), bilateral vestibulopathy (BVP), and neuropathy among 82 patients with DBN carrying an *FGF14* (GAA)_≥250_ expansion.

The frequency of *FGF14* (GAA)_≥250_ expansions in DBN syndromes stratified by phenotype was 56% (42/75) for DBN plus additional isolated cerebellar ocular motor signs, 72% (23/32) for DBN plus cerebellar ataxia, 30% (17/56) for DBN plus cerebellar ocular motor signs and/or ataxia and extracerebellar features, which included BVP and/or polyneuropathy, and 0% (0/7) for pure DBN (**Fig. 2B**).

### Phenotype and Discriminative Features of (GAA)_≥250_-*FGF14*-related Downbeat Nystagmus Syndromes

Table 1 summarizes the baseline characteristics and clinical features of patients carrying a (GAA)_≥250_ expansion, a (GAA)_200-249_ allele, and a (GAA)_<200_ allele.

**Table 1:** Characteristics and discriminative features of GAA-*FGF14*-related DBN syndromes. Unless specified, data are reported as median (range). Bold values indicate statistically significant adjusted *p* values (*q* values) below the 0.05 threshold as per the Benjamini-Hochberg method. Data on age at onset were missing for eight patients in the (GAA)_≥250_ group, three patients in the (GAA)_200-249_ group, and 10 patients in the (GAA)_<200_ group. *The *FGF14* (GAA)_≥250_ group includes a patient carrying biallelic GAA repeat expansions (280 and 313 repeat units). The *FGF14* (GAA)_<200_ group includes two patients carrying a likely non-pathogenic (GAAGGA)_n_ expansion (335 and 319 triplet repeat units equivalent, respectively). **In patients without bilateral vestibulopathy. Abbreviations: BVP: bilateral vestibulopathy; FARS: Friedreich Ataxia Rating Scale; vHIT: video head impulse test; VOR: vestibulo-ocular reflex.

|  | *FGF14* (GAA)<sub>≥250</sub><br>(n=82) | *FGF14* (GAA)<sub>200-249</sub><br>(n=20) | *FGF14* (GAA)<sub><200</sub><br>(n=68) | *FGF14* (GAA)<sub>≥250</sub> vs<br>*FGF14* (GAA)<sub><200</sub> |  |
| --- | --- | --- | --- | --- | --- |
|  |  |  |  | *p* value | *q* value |
| Male sex | 44 (54%) | 11 (55%) | 40 (59%) | - | - |
| Triplet repeat count of the larger allele* | 324 (250-516) | 234 (207-249) | 37 (8-335) | - | - |
| Age at disease onset | 67 (30-81) | 72 (45-84) | 66 (17-88) | 0.611 | 0.760 |
| Disease duration | 6 (0-26.5) | 4 (0-23) | 4 (0-50) | 0.180 | 0.407 |
| Age at last examination | 73 (40-92) | 79 (58-86) | 72 (21-91) | 0.346 | 0.554 |
| Positive family history | 31/82 (38%) | 4/20 (20%) | 13/67 (19%) | 0.019 | 0.097 |
| FARS disability stage | 3 (1.5-5) | 3 (1.5-4) | 2 (1-4) | 0.115 | 0.307 |
| History of falls | 27/50 (54%) | 8/14 (57%) | 20/35 (57%) | 0.827 | 0.882 |
| Regular use of walking aid | 16/81 (20%) | 4/19 (21%) | 15/66 (23%) | 0.688 | 0.760 |
| **Symptoms** |  |  |  |  |  |
| Episodic symptoms | 11/81 (14%) | 0/19 (0%) | 18/66 (27%) | 0.059 | 0.173 |
| Gait impairment | 81/81 (100%) | 20/20 (100%) | 68/68 (100%) | 1.000 | 1.000 |
| Vertigo or dizziness | 23/82 (28%) | 10/20 (50%) | 26/68 (38%) | 0.222 | 0.442 |
| Visual disturbances | 47/82 (57%) | 8/20 (40%) | 30/68 (44%) | 0.140 | 0.344 |
| Fine motor impairment | 12/80 (15%) | 1/18 (6%) | 12/66 (18%) | 0.658 | 0.760 |
| Speech impairment | 13/81 (16%) | 4/19 (21%) | 13/66 (20%) | 0.665 | 0.760 |
| **Clinical signs** |  |  |  |  |  |
| Impaired balance/gait | 67/79 (85%) | 13/19 (68%) | 44/64 (69%) | 0.027 | 0.108 |
| **Cerebellar ocular motor signs** |  |  |  |  |  |
| - Gaze-evoked nystagmus | 57/82 (70%) | 11/20 (55%) | 42/67 (63%) | 0.390 | 0.594 |
| - Saccadic pursuit | 80/81 (99%) | 20/20 (100%) | 59/67 (88%) | 0.011 | 0.073 |
| - Dysmetric saccades | 22/79 (28%) | 6/20 (30%) | 22/67 (33%) | 0.588 | 0.760 |
| - Impaired VOR suppression** | 64/74 (86%) | 16/17 (94%) | 23/46 (50%) | <0.001 | **<0.001** |
| **Cerebellar ataxia** |  |  |  |  |  |
| - Ataxia of upper limbs | 16/73 (22%) | 1/14 (7%) | 21/55 (38%) | 0.051 | 0.163 |
| - Dysidiadochokinesia | 15/71 (21%) | 1/16 (6%) | 14/54 (26%) | 0.531 | 0.760 |
| - Dysarthria | 12/80 (15%) | 2/19 (11%) | 12/63 (19%) | 0.653 | 0.760 |
| Tremor of upper limbs | 12/79 (15%) | 2/18 (11%) | 6/66 (9%) | 0.318 | 0.536 |
| **Neuropathy** |  |  |  |  |  |
| - Impaired vibration at ankle (≤3/8) | 11/79 (14%) | 6/18 (33%) | 24/65 (37%) | 0.002 | **0.018** |
| - Ankle hyporeflexia | 20/79 (25%) | 4/18 (22%) | 29/65 (45%) | 0.021 | 0.097 |
| Pyramidal tract signs | 1/77 (1%) | 0/18 (0%) | 2/66 (3%) | 0.595 | 0.760 |
| **MRI** |  |  |  |  |  |
| Vermis atrophy | 8/60 (13%) | 1/11 (9%) | 10/47 (21%) | 0.307 | 0.536 |
| Cerebellar hemisphere atrophy | 5/60 (8%) | 0/11 (0%) | 8/47 (17%) | 0.235 | 0.442 |
| Supratentorial atrophy | 5/60 (8%) | 2/10 (20%) | 4/47 (9%) | 1.000 | 1.000 |
| Brainstem atrophy | 0/60 (0%) | 0/10 (0%) | 2/47 (4%) | 0.191 | 0.407 |
| **Nerve conduction studies** |  |  |  |  |  |
| Axonal neuropathy | 7/15 (47%) | 3/5 (60%) | 10/11 (91%) | 0.036 | 0.129 |
| **Vestibular function evaluation – caloric stimulation, vHIT, rotatory chair** |  |  |  |  |  |
| Bilateral vestibulopathy | 8/79 (10%) | 3/18 (17%) | 20/62 (32%) | 0.001 | **0.018** |
| VOR gain on vHIT in BVP – mean (±SD) | 0.44 (±0.10) | 0.21 | 0.18 (±0.17) | 0.006 | **0.0496** |

Symptoms in (GAA)_≥250_-*FGF14* patients with DBN started at a median age of 67 years (range, 30-81). The most common presenting symptom was gait unsteadiness (89%; 72/81), followed by visual disturbances (37%; 30/81), such as diplopia, oscillopsia, or visual blurring, vertigo and/or dizziness (20%; 16/81), and, rarely, dysarthria (1%; 1/81) (**Fig. 2C**). We observed a weak inverse correlation between the age at onset and the size of the repeat expansion (74 patients; Pearson’s r, -0.30; R^2^=0.09; *p*=0.01) (**Fig. 2D**). Family history for DBN or ataxia was positive in 31 patients (38%), of whom 26 had evidence of autosomal dominant inheritance. In keeping with the likely reduced male transmission of the disease due to contraction of the *FGF14* repeat locus upon paternal transmission,^12,25^ we observed that only 2 of 26 dominantly inherited cases (8%) were paternally inherited.

The median age and disease duration at last examination were 73 years (range, 40-92) and 6 years (range, 0-26.5), respectively. Additional cerebellar ocular motor signs were observed in 100% (82/82), cerebellar ataxia in 43% (35/82), and extracerebellar features in 21% (17/82) of (GAA)_≥250_-*FGF14* patients (**Fig. 2E**). Cerebellar ocular motor signs comprised – in decreasing order of frequency – saccadic pursuit (99%; 80/81), impaired visual fixation suppression of the VOR (86%; 64/74 in patients without BVP), gaze-evoked nystagmus (70%; 57/82), dysmetric saccades (28%; 22/79), and rebound nystagmus (20%; 16/82). Signs of cerebellar ataxia included dysmetria of upper limbs (46%; 16/35), dysdiadochokinesia (47%; 15/32), dysarthria (35%; 12/34), and intention tremor (11%; 4/35). Cerebellar involvement reflected by ∼3Hz titubation was identified in 18 of 55 patients (33%) on posturography. Brain MRI of eight patients showed cerebellar atrophy (13%; 8/60), which was limited to the vermis in three patients and extended to the hemispheres in five patients. Two patients with discrete pan-cerebellar atrophy showed no signs of ataxia on examination.

The subgroup of (GAA)_≥250_-*FGF14* patients with DBN plus cerebellar ocular motor signs and/or ataxia and extracerebellar features comprised nine patients with polyneuropathy, six patients with BVP, and two patients with polyneuropathy plus BVP (**Fig. 2E**). Polyneuropathy and BVP occurred more frequently in patients with cerebellar ataxia (71%; 12/17) compared to patients exhibiting only cerebellar ocular motor signs (29%; 5/17) (**Fig. 2E**). Polyneuropathy was diagnosed on nerve conduction studies in seven patients and clinically in four patients. Results of nerve conduction studies were consistent with length-dependent sensorimotor axonal neuropathy in six patients and sensory neuropathy in one patient. BVP was diagnosed in 8 of 79 patients (10%) by caloric stimulation (*n*=1) and vHIT (*n*=7). Disease duration at time of vestibular assessment differed significantly between patients with and without BVP (median, 14 vs 4 years; Mann-Whitney U test, *p*=0.0018).

To identify discriminative features of DBN syndromes in (GAA)_≥250_-*FGF14* patients, we compared their phenotypic features to that of (GAA)_<200_-*FGF14* patients (**Table 1**). Both groups did not differ significantly in terms of disease duration, baseline characteristics, functional impairment, and symptoms. Impairment of the visual fixation suppression of the VOR was strongly associated with (GAA)_≥250_-*FGF14* DBN syndromes (86% vs 50%; OR, 6.29; 95% CI, 2.45-17.24; Fisher’s exact test, *p*=2.089e-05, *q*=6.685e-04). In keeping with the lower frequency of *FGF14* repeat expansions in patients with DBN plus extracerebellar features, impaired vibration at the ankles (14% vs 37%; OR, 0.28; 95% CI, 0.11-0.66; Fisher’s exact test, *p*=0.002, *q*=0.018) and BVP (10% vs 32%; OR, 0.24; 95% CI, 0.08-0.63; Fisher’s exact test, *p*=0.001, *q*=0.018) were significantly less common in (GAA)_≥250_-*FGF14* than in (GAA)_<200_-*FGF14* patients. Furthermore, in the subset of patients in whom BVP was objectified on vHIT, the mean VOR gain was significantly higher (values of >0.6 are considered normal) in (GAA)_≥250_-*FGF14* compared to (GAA)_<200_-*FGF14* patients (0.44 ±0.10 vs 0.18 ±0.17; Mann-Whitney U test, *p*=0.006, *q*=0.0496), despite no significant difference in disease duration between the groups (median, 14.5 vs 8 years; Mann-Whitney U test, *p*=0.1512). These results suggest that BVP occurs late in the course of (GAA)_≥250_-*FGF14*-related DBN syndromes and remains mild.

### Disease Progression in (GAA)_≥250_-*FGF14*-related Downbeat Nystagmus Syndromes

We used a Kaplan-Meier survival analysis to model the age at disease onset of (GAA)_≥250_-*FGF14* patients, estimating their probability of remaining disease-free over time (**Fig. 3A**). Survival until disease onset in (GAA)_≥250_-*FGF14* patients did not significantly differ between the three main phenotypic subgroups (median age at onset, 67 years [33-80] for DBN+COM vs 67 years [36-81] for DBN+CA vs 66 years [30-78] for DBN+EC; Cox-Mantel test, *X^2^*(2)=0.1267; *p*=0.939). However, disease duration at last examination was significantly longer in patients with DBN plus cerebellar and extracerebellar features (median, 4.25 years [0-16] for DBN+COM vs 6 years [1-22] for DBN+CA vs 9 years [3-26.5] for DBN+EC; Kruskal-Wallis test, *p*=0.014) (**Fig. 3B**), suggesting that extracerebellar features may develop later in the disease course.

**Figure 3:**
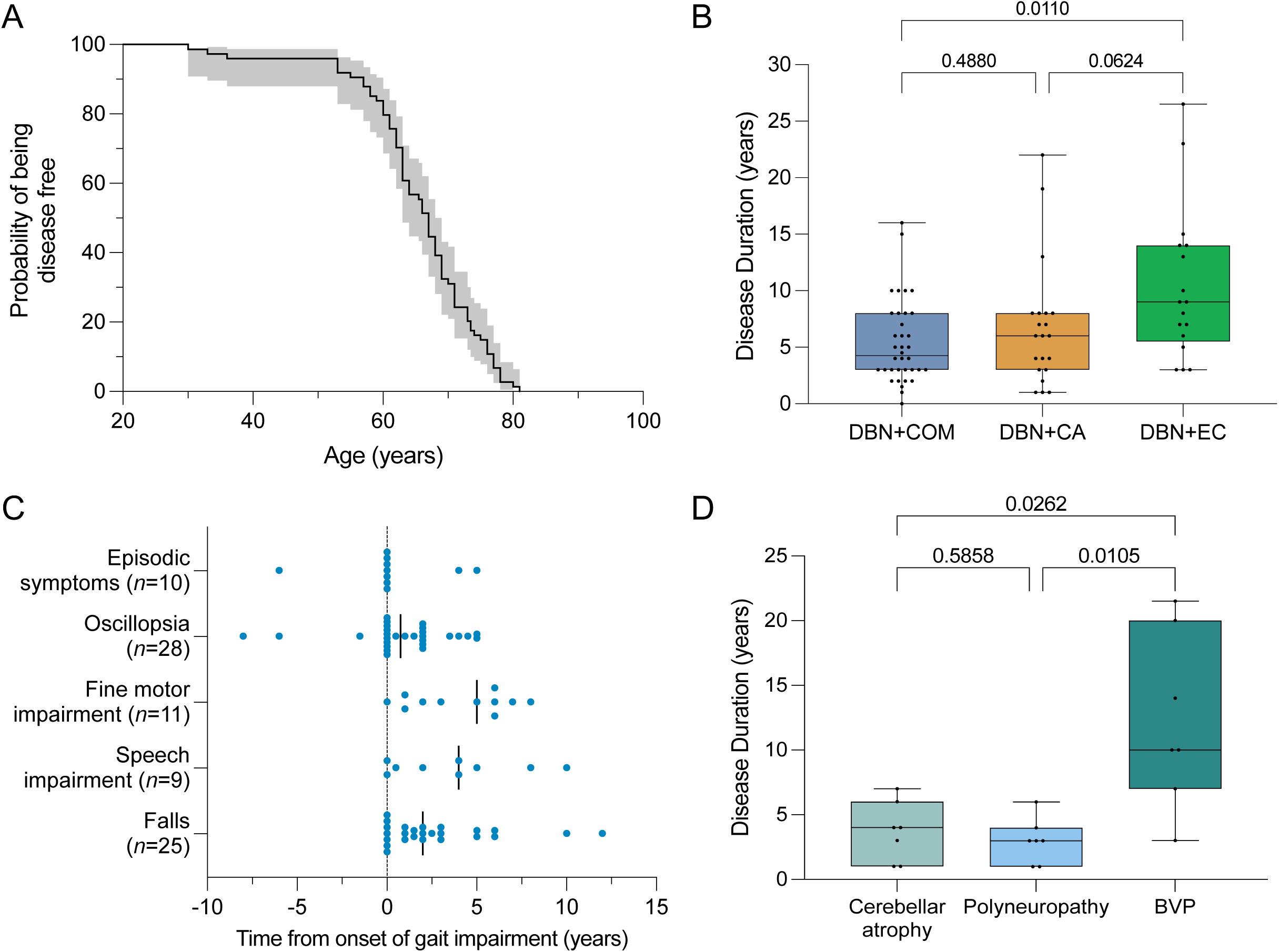
Phenotypic evolution in GAA-*FGF14*-related DBN syndromes. (A) Kaplan-Meier curve showing the probability of being disease free as a function of the age (years) for 74 patients carrying an *FGF14* (GAA)_≥250_ expansion for whom age at onset was available. Median survival before occurrence of disease was 67 years. The shaded gray area displays the 95% confidence interval around the probability estimate. (B) Disease duration at last examination for patients carrying an *FGF14* (GAA)_≥250_ expansion with DBN plus additional isolated cerebellar ocular motor signs (DBN+COM, *n*=36; median, 4.25 years), DBN plus cerebellar ataxia (DBN+CA, *n*=21; median, 6 years), and DBN plus cerebellar ocular motor signs and/or ataxia and extracerebellar features (DBN+EC, *n*=17; median, 9 years). Disease duration was significantly longer for patients with extracerebellar features (Kruskal-Wallis test, *p*=0.014). (**C**) Temporal evolution of select phenotypic features relative to the onset of gait impairment (dotted line). The solid black lines show the median time from onset of gait impairment for each individual feature. (**D**) Occurrence of cerebellar atrophy on brain MRI (*n*=7), polyneuropathy on nerve conduction studies (*n*=7), and BVP on caloric stimulation or vHIT (*n*=7) in relation to disease onset. BVP occurred later in disease course compared to cerebellar ataxia and polyneuropathy (Kruskal-Wallis test, *p*=0.004). In panels B and D, the individual between-group adjusted *p* values (*q* values) as per the Benjamini-Hochberg method are shown in the graphs. Adjusted *p* values < 0.05 indicate statistically significant difference.

We also studied the evolution of key symptoms in (GAA)_≥250_-*FGF14* patients in relation to gait impairment (**Fig. 3C**). Episodic symptoms (median, 0 year; range, -6-5) and oscillopsia (0.75 year; -8-5) manifested concurrently with gait impairment in most patients. However, episodic symptoms and oscillopsia preceded gait impairment in 10% (1/10) and 11% (3/28) of patients for whom information on age at onset of these features was available, respectively. In comparison, fine motor impairment (5 years; 0-8), speech impairment (4 years; 0-10), and falls (2 years; 0-12) developed later in the disease course, consistent with the findings from a recent progression study.^14^ Sixteen patients (20%) eventually required a walking aid after a median disease duration of 4 years (range, 0-13), including three patients (4%) who became wheelchair dependent after a median disease duration of 8 years (range, 7-12). Further to this analysis, we observed that BVP developed at a significantly later stage in the disease compared to cerebellar atrophy and neuropathy (median disease duration at first occurrence, 4 years [1-7] for cerebellar atrophy vs 3 years [1-6] for polyneuropathy vs 10 years [3-21.5] for BVP; Kruskal-Wallis test, *p*=0.004) (**Fig. 3D**).

The progression of functional disability in (GAA)_≥250_-*FGF14* patients was assessed by means of the FARS functional staging (**Fig. 4**). At time of last examination, the median FARS stage measured off 4-AP was 3 (range, 2-5), indicating a mild level of disability. We found no association between the FARS functional stage and disease duration (73 patients; Spearman’s rho, 0.194; *p*=0.10) (**Fig. 4A**). Neither age, disease duration, or expansion size was associated with the cross-sectional FARS stage in a multiple linear regression model (*F*(3,69)=2.602, adjusted R^2^=0.06, *p*=0.059; age, b=0.017, *p*=0.111; disease duration, b=0.027, *p*=0.151; expansion size, b=-0.001, *p*=0.527). Analysis of the cross-sectional and longitudinal FARS stage data revealed that half of patients (27/54) displayed at least mild disability within the first five years of disease onset. However, disability remained at most mild in the majority of patients (86%, 63/73) even after more than 25 years of disease duration. This observation was further supported by longitudinal data in 40 patients (148 observations) showing an overall slow intra-individual increase of 0.10 FARS stage per year of disease (standard error, 0.02 by linear mixed-effects model) (**Fig. 4B**). While disease evolved relatively slowly in most patients, inspection of individual disability progression trajectories revealed inter-individual variability in rates of progression (**Fig. 4B**). Concurrent medical conditions substantially contributed to disability burden in two patients. One patient became wheelchair-bound following a complicated medical admission. However, this patient also carried biallelic expansions (280 and 313 repeat units) which may have contributed to the faster disability progression. The second patient, carrying alleles of 218 and 270 repeat units, became wheelchair-bound following a hip fracture.

**Figure 4:**
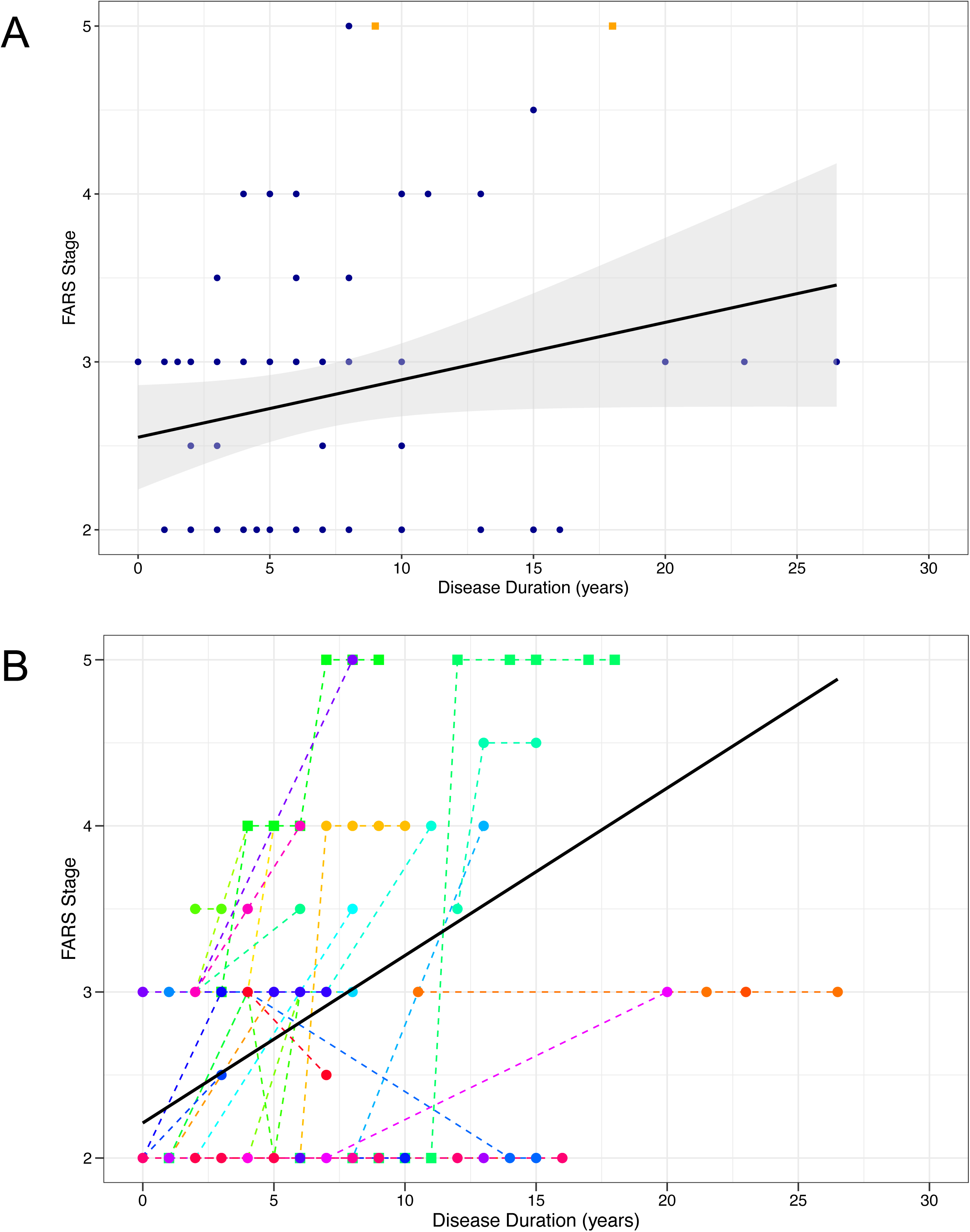
Progression of disability in GAA-*FGF14*-related DBN syndromes. (A) Cross-sectional progression of the functional impairment as assessed by the FARS functional disability stage relative to disease duration (*n*=73 patients carrying an *FGF14* [GAA]_≥250_ expansion). The FARS functional stage was not significantly associated with disease duration (Spearman’s rho, 0.194; *p*=0.10). (**B**) Longitudinal intra-individual progression of functional impairment as assessed by the FARS functional disability stage relative to disease duration (148 observations from 40 patients carrying an *FGF14* [GAA]_≥250_ expansion are shown). Observations from the same patient are connected by a dotted line. The solid black line shows the average progression of the FARS stage over disease duration across all patients as modelled by a linear mixed-effects model accounting for disease duration as fixed effect and with random intercepts and random slopes. In both panels, only FARS stages measured off 4-AP treatment are shown. Two patients who became wheelchair-dependent following a hip fracture and a prolonged medical admission, respectively, are represented by orange squares in panel A and by squares in panel B. These patients also carried alleles of 218 and 270 repeat units and 280 and 313 repeat units, respectively.

### (GAA)_200-249_-*FGF14* Patients have a Similar Clinical Phenotype and Disease Evolution as (GAA)_≥250_-*FGF14* Patients

While previous studies have established a preliminary pathogenic threshold of at least 250 GAA repeat units,^12,13^ the pathogenic potential of (GAA)_200-249_ alleles has recently been suggested following the identification of a family with late-onset slowly progressive ataxia in which (GAA)_200-249_ alleles and (GAA)_≥250_ expanded alleles segregated with disease.^24^ Alleles of 200 to 249 GAA repeat units were significantly enriched in patients with DBN compared to controls (12% vs 0.87%; OR, 15.20; 95% CI, 7.52-30.80; Fisher’s exact test, *p*=9.876e-14), thus providing further support for their potential pathogenicity.

We next aimed to characterize the phenotype of (GAA)_200-249_-*FGF14* patients to assess whether it resembled that of (GAA)_≥250_-*FGF14* patients (**Table 1**). (GAA)_200-249_-*FGF14* patients did not significantly differ from (GAA)_≥250_-*FGF14* patients in terms of baseline characteristics, symptoms, clinical signs, and progression of functional disability. In comparison, impairment of the visual fixation suppression of the VOR (indicating an involvement of the fixation system), which was strongly associated with (GAA)_≥250_-*FGF14*-related DBN syndromes, was significantly more common in (GAA)_200-249_-*FGF14* patients compared to (GAA)_<200_-*FGF14* patients (94%, 16/17 vs 50%, 23/46; OR, 15.45; 95% CI, 2.07-696.97; Fisher’s exact test, *p*=0.001; *q*=0.037), but not different from (GAA)_≥250_-*FGF14* patients (94%, 16/17 vs 86%, 64/74; Fisher’s exact test, *p*=0.682; *q*=1.000).

### Association of the rs72665334 Variant with *FGF14* (GAA)_≥250_ Expansions in Patients with Downbeat Nystagmus

A recent GWAS has identified an association between DBN and the *FGF14* rs72665334 C>T variant in 106 patients (**Fig. 5A**).^11^ This same variant, which has an allele frequency of ∼7% in the European population (1000 Genomes Project^27^), was also found to be part of a disease haplotype shared by three Australian patients with GAA-*FGF14* ataxia in a previous study.^13^ Given the frequent occurrence of DBN in GAA-*FGF14* ataxia, we therefore hypothesized that the rs72665334 variant might be in disequilibrium with the *FGF14* GAA repeat expansion, which is located ∼11kb away (**Fig. 5A)**. In the DBN cohort, we observed that a significantly greater proportion of patients with a C|T or T|T genotype carried an *FGF14* (GAA)_≥250_ expansion compared to an *FGF14* (GAA)_<200_ allele (54%, 19/35 vs 14%, 4/29; OR, 7.18; 95% CI, 1.91 to 34.43; Fisher’s exact test *p*=0.0014) (**Fig. 5B**). Similarly, we also found that 38% of patients (14/37) from an independent and ethnically distinct cohort of 37 French-Canadian index patients with GAA-*FGF14* ataxia and DBN had a C|T or T|T rs72665334 genotype. To assesses whether the rs72665334 variant is in disequilibrium with the *FGF14* (GAA)_≥250_ expansion at a population level, we compared the frequency of the C|T and T|T genotypes in 503 European controls from the 1000 Genomes Project and 72 patients with GAA-*FGF14* ataxia and DBN (35 patients from this study and 37 French-Canadian patients). The C|T and T|T genotypes were significantly more frequent in patients with DBN carrying an *FGF14* (GAA)_≥250_ expansion compared to controls (46%, 33/72 vs 12%, 60/503; OR, 6.22; 95% CI, 3.51 to 11.02; Fisher’s exact test, *p*=1.063e-10) (**Fig. 5C**). Together, these results show that the rs72665334 variant is in disequilibrium with the *FGF14* GAA repeat expansion. However, its absence in 54% of (GAA)_≥250_-*FGF14* patients suggests that *FGF14* GAA repeat expansions may arise on distinct haplotype backgrounds.

**Figure 5:**
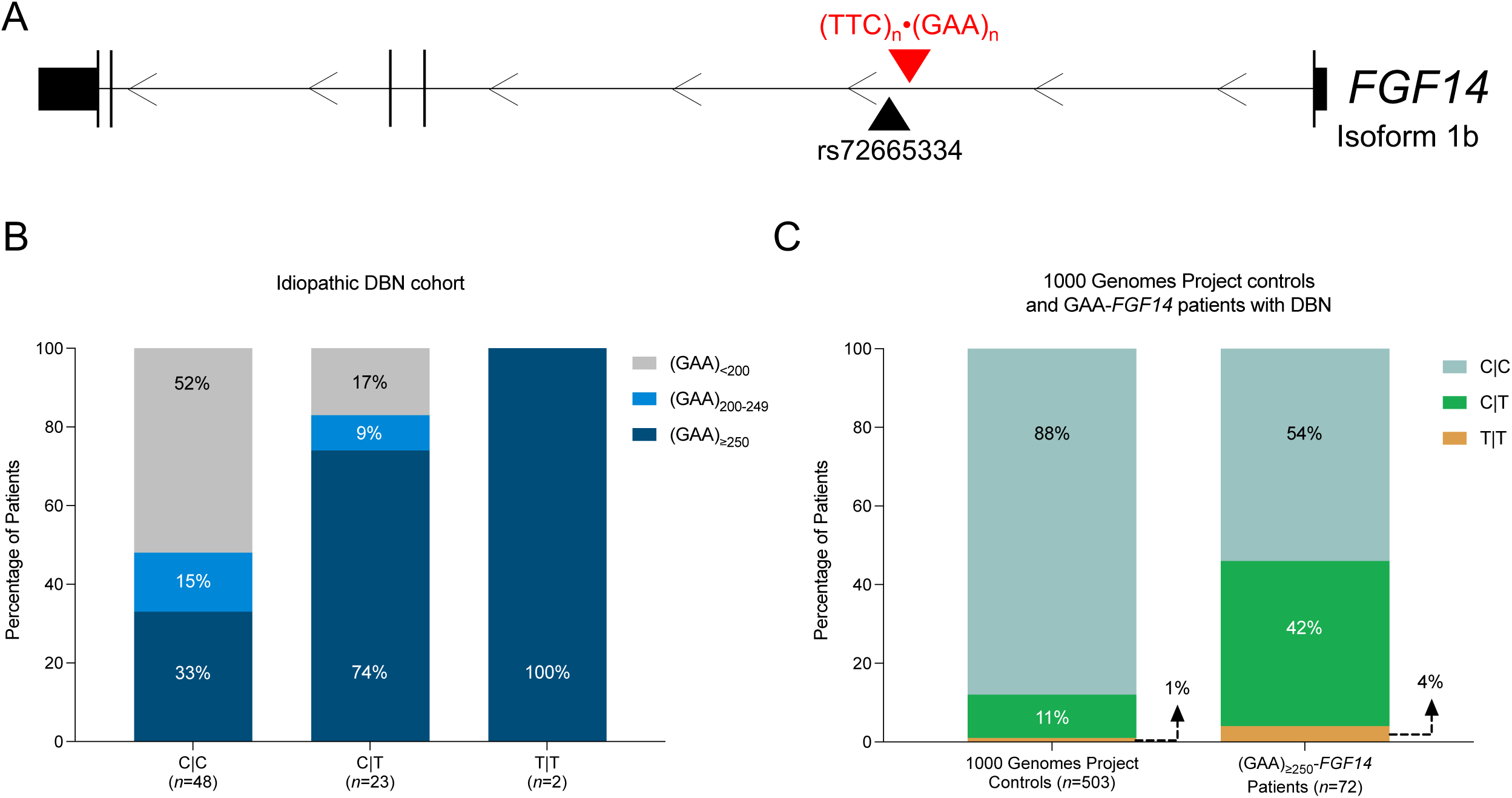
Association of the rs72665334 variant with *FGF14* GAA expansions. (**A**) Diagram of the *FGF14* gene, isoform 1b showing the location of the rs72665334 variant (GRCh38, chr13:102,150,076) in relation to the (GAA)_n_•(TTC)_n_ repeat locus in the first intron (GRCh38, chr13:102,161,575-102,161,726). The rs72665334 variant is located ∼11kb 5’ of the intronic GAA short tandem repeat. (**B**) Percentage of patients with DBN carrying an *FGF14* (GAA)_≥250_ expansion (*n*=35; dark blue), an *FGF14* (GAA)_200-249_ allele (*n*=9; light blue), and an *FGF14* (GAA)_<200_ allele (*n*=29; gray) with the C|C, C|T, and T|T rs72665334 genotypes. There was a significantly greater proportion of patients with a C|T or T|T genotype who carried an *FGF14* (GAA)_≥250_ expansion compared to an *FGF14* (GAA)_<200_ allele (54% vs 14%; OR, 7.18; 95% CI, 1.91 to 34.43; Fisher’s exact test *p*=0.0014). (**C**) Frequency of the C|C, C|T, and T|T rs72665334 genotypes in 503 European controls from the 1000 Genomes Project^27^ and 72 patients with DBN carrying an *FGF14* (GAA) expansion (*n*=35 from the idiopathic DBN cohort and *n*=37 from the French-Canadian cohort). The C|T and T|T genotypes were significantly more frequent in patients with DBN carrying an *FGF14* (GAA)_≥250_ expansion compared to controls (46%, 33/72 vs 12%, 60/503; OR, 6.22; 95% CI, 3.51 to 11.02; Fisher’s exact test, *p*=1.063e-10).

### Treatment Response to 4-Aminopyridine

#### Open-label real-world treatment response data

We assessed the response to 4-AP treatment in patients with DBN stratified by *FGF14* genotype (open-label treatment as part of routine clinical care). 81% of (GAA)_≥250_-*FGF14* patients (66/81), 61% of (GAA)_200-249_-*FGF14* patients (11/18), and 56% of (GAA)_<200_-*FGF14* patients (37/66) had previously received 4-AP. Treatment response for all patients had been recorded before the discovery of GAA-*FGF14* ataxia and, therefore, both clinicians and patients were naturally blind to the underlying GAA-*FGF14* genotype (“real-world double-blind” to genotype). A clinician-reported treatment response was recorded for 81% of (GAA)_≥250_-*FGF14* patients (29/36; improvement of ocular motor signs, including DBN: 24 patients; and gait: 14 patients), 80% of (GAA)_200-249_-*FGF14* (4/5; ocular motor signs: 2; gait: 3), and 31% of (GAA)_<200_-*FGF14* patients (5/16; ocular motor signs: 3; gait: 2) (**Fig. 6A**). A patient-reported benefit was recorded for 59% of (GAA)_≥250_-*FGF14* (29/49; improvement of visual symptoms: 14 patients; and gait: 17 patients), 60% of (GAA)_200-249_-*FGF14* (3/5; visual symptoms: 2; gait: 2), and 11% of (GAA)_<200_-*FGF14* patients (2/19; visual symptoms: 1; gait: 1) (**Fig. 6B**). (GAA)_≥250_-*FGF14* and (GAA)_200-249_-*FGF14* patients had a significantly greater clinician-reported (80% vs 31%; OR, 8.63; 95% CI, 2.08 to 41.96; Fisher’s exact test *p*=0.0011) and patient-reported (59% vs 11%; OR, 11.96; 95% CI, 2.45 to 117.27; Fisher’s exact test *p*=0.0003) response rate to 4-AP treatment compared to (GAA)_<200_-*FGF14* patients. To next evaluate the effect of 4-AP on disability, we examined the FARS disability stages assessed on and off 4-AP. Data were available for seven (GAA)_≥250_-*FGF14* and two (GAA)_200-249_-*FGF14* patients (**Fig. 7**). Remarkably, the FARS stages for all nine patients were invariably lower, by up to 2 stages, while on 4-AP compared to off 4-AP. The therapeutic benefit of 4-AP was further evidenced by the clear association between drug intake and improvement of disability in four patients in whom stopping the drug led to an increase in disability (**Fig. 7**). Given the average progression of 0.10 FARS stage per year of disease (**Fig. 4B**), an improvement of 1 FARS stage on 4-AP treatment might be considered equivalent to “reversing” the disease progression by approximately 10 years. These results, which need confirmation in randomized placebo-controlled trials, suggest that the GAA-*FGF14* genotype status stratifies a subgroup of patients with DBN who are high responders to 4-AP. The response to 4-AP, whether reported by the clinician or the patient, may also be a useful clinical sign to increase suspicion for GAA-*FGF14* disease, given its positive predictive value of 88% (95% CI, 74-96%) for (GAA)_≥250_-*FGF14* status and 89% (95% CI, 76-96%) for combined (GAA)_≥250_-*FGF14* and (GAA)_200-249_-*FGF14* statuses.

**Figure 6:**
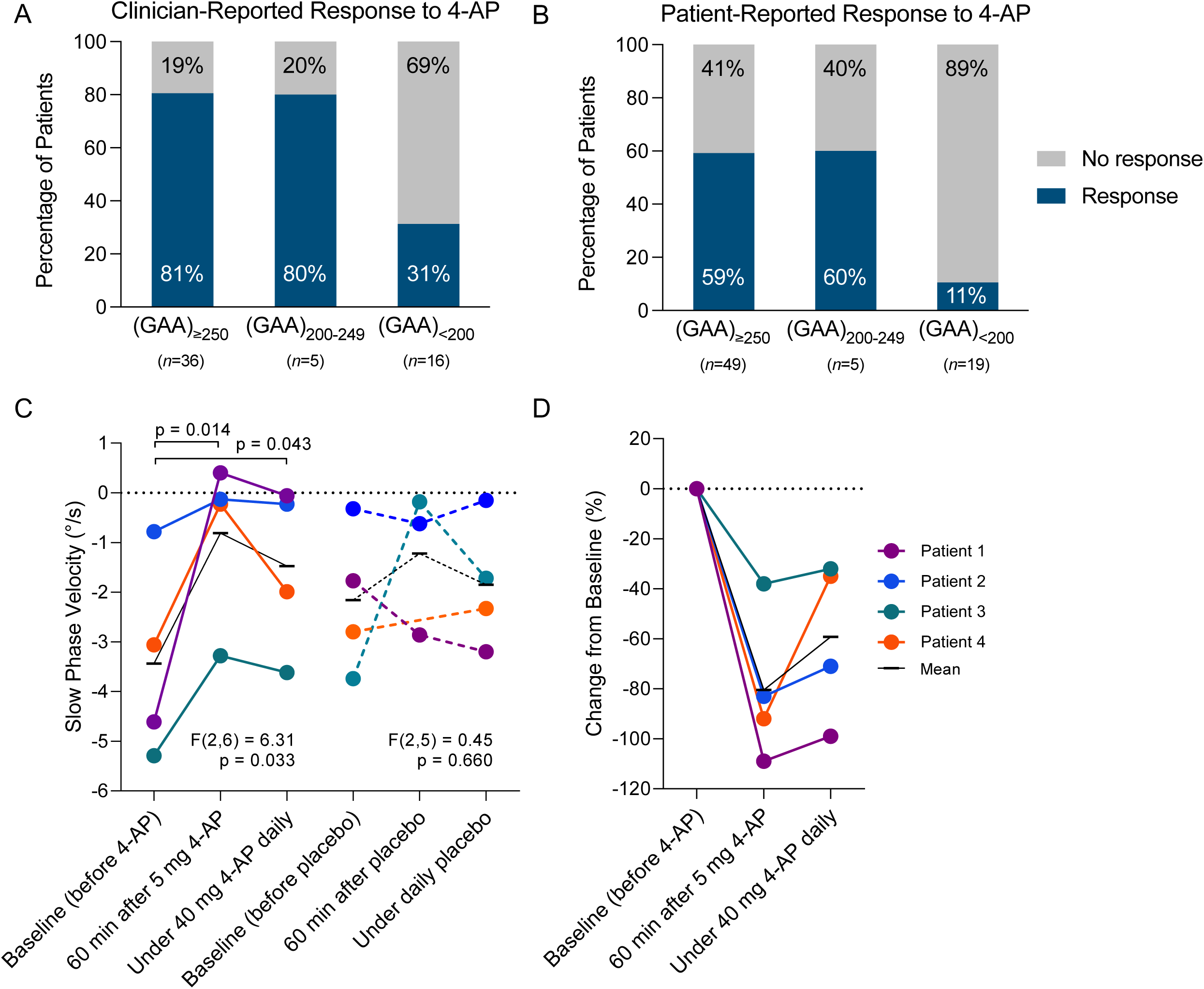
Treatment response to 4-aminopyridine. *Open-label real-world treatment response data*. Percentage of patients presenting (**A**) a clinician-reported response and (**B**) a patient-reported response to 4-AP treatment in the subgroups with an *FGF14* (GAA)_≥250_ expansion (29/36 and 29/49 patients, respectively), (GAA)_200-249_ allele (4/5 and 3/5 patients), and (GAA)_<200_ allele (5/16 and 2/19 patients). *Double-blind placebo-controlled clinical trial data*. (**C**) Effect of 4-AP on the slow phase velocity (SPV) of DBN in a randomized double-blind trial.^16^ Compared to baseline, the absolute value of the SPV significantly decreased under 4-AP 60 min after the first dose and under daily treatment, but not under placebo. Note that all four patients were randomized to receive active treatment first (= 4-AP) and placebo second. SPV during all three measurements of the placebo phase was higher than at the baseline of the treatment phase. The improved values of the SPV under 4-AP were in the range of the placebo values. (**D**) Treatment effect of 4-AP, illustrated by relative reduction (improvement) of SPV as potential future trial outcome assessment. The genotype of the four patients was: patient 1, 65 and 298 repeat units; patient 2, 44 and 354 repeat units; patient 3, 16 and 265 repeat units; patient 4, 280 and 313 repeat units. In panels C and D, the black line shows the mean value for all four patients at each time point.

**Figure 7:**
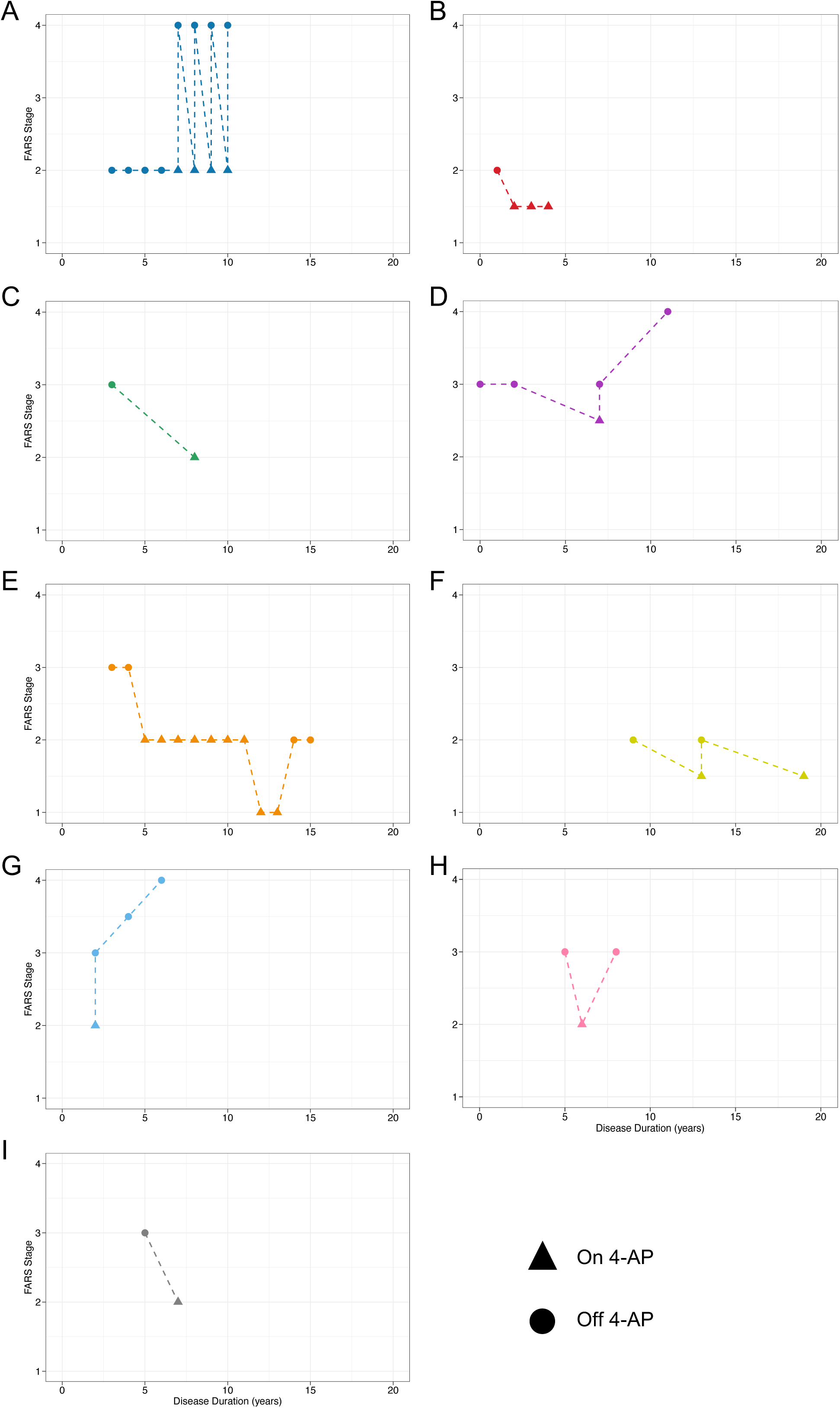
Effect of 4-aminopyridine on functional impairment. Longitudinal intra-individual progression of functional impairment while on and off 4-AP treatment as assessed by the FARS functional disability stage relative to disease duration for (**A-G**) seven patients carrying an *FGF14* (GAA)_≥250_ expansion and (**H-I**) two patients carrying an *FGF14* (GAA)_200-249_ allele. FARS stages were invariably lower while on 4-AP treatment for all nine patients, indicating an improvement in the level of disability. No patients carrying an *FGF14* (GAA)_<200_ allele had FARS stages recorded while on and off 4-AP treatment.

#### Double-blind placebo-controlled clinical trial data

We next analyzed in a first preliminary probing approach whether the effects of 4-AP treatment in GAA-*FGF14*-related DBN could be validated not only in open-label assessments, but also in double-blind, placebo-controlled randomized treatment assessments. For this purpose, we re-analyzed placebo-controlled video-oculography data of four patients with DBN – now known to be (GAA)_≥250_-*FGF14* – from a previous randomized double-blind trial.^16^ In addition to being double-blind, all trial assessments were blinded to the underlying GAA-*FGF14* genotype, which was not discovered until after the trial. The absolute value of the SPV decreased under treatment with 4-AP (*F*(2,6) = 6.31, *p*=0.033), but not under placebo (*F*(2,5) = 0.45, *p*=0.660) (**Fig. 6C**). The decrease in SPV was significant 60 min after administration of a single dose of 5 mg of 4-AP compared to intra-individual baseline measurements (least squares mean difference: 2.63°/s, *p*=0.014), and persisted under a stable dose of 40 mg of 4-AP daily (least squares mean difference: 1.97°/s, *p*=0.043). This effect corresponds to relative reductions (improvements) of SPV by 38-109% after a single dose, and 32-99% under stable treatment with 4-AP (**Fig. 6D**).

## Discussion

This study tested the hypothesis that *FGF14* (GAA)_≥250_ repeat expansions are a recurrent cause of idiopathic DBN syndromes, where DBN may be the presenting phenotype of a wider GAA-*FGF14*-related neurological disease spectrum. Our findings identify *FGF14* (GAA)_≥250_ repeat expansions as a highly frequent monogenic cause of DBN, particularly when associated with additional cerebellar features. Through systematic in-depth phenotyping, we characterized the disease evolution of (GAA)_≥250_-*FGF14*-related DBN and its treatment response to 4-AP. Additionally, we explored the phenotypic and 4-AP treatment response characteristics of patients with *FGF14* (GAA)_200-249_ alleles, whose pathogenicity has recently been suggested.

The cause of DBN remains unknown in approximately 30% of cases (idiopathic DBN).^1,2^ Identifying genetic causes for these cases has proven challenging. We now show that *FGF14* (GAA)_≥250_ repeat expansions account for almost 50% of cases of previously unexplained DBN in a large and strictly consecutive cohort of European patients. Dominantly inherited *FGF14* GAA repeat expansions have recently been shown to be a common cause of spinocerebellar ataxia (SCA27B), which is associated with DBN in up to 40-60% of patients.^12,14^ However, the high frequency of *FGF14* (GAA)_≥250_ repeat expansions in patients with a predominantly sporadic non-ataxic DBN presentation was unexpected. One explanation may be that DBN is a milder phenotypic presentation of GAA-*FGF14* disease, in which overt cerebellar ataxia and other multisystemic involvement can be absent (or, in some cases, develop later in the disease course). Our results further suggest that genetic testing for *FGF14* GAA repeat expansion should now become part of the diagnostic work-up of patients with idiopathic DBN.

By screening a cohort of patients with idiopathic DBN, our study closes a gap in the delineation of the phenotypic spectrum and evolution of GAA-*FGF14* disease. The identification of additional floccular/parafloccular cerebellar ocular motor signs in all (GAA)_≥250_-*FGF14* patients with DBN indicates that the basic dysfunction arises from this cerebellar region and that pure DBN is an uncommon manifestation in GAA-*FGF14* disease. Furthermore, the observation of DBN and ocular motor signs manifesting up to 8 years prior to the development of gait impairment suggests that such signs may present in isolation early in the disease course. However, the recurrent identification of *FGF14* expansions in patients with DBN plus additional isolated cerebellar ocular motor signs but without overt ataxia despite disease duration of up to 16 years raises the possibility that GAA-*FGF14* disease may remain limited to the cerebellar ocular motor system without broader cerebellar involvement in a subset of patients. This indicates that cerebellar ataxia is not a universal and mandatory feature of GAA-*FGF14* disease and that ocular motor signs and ataxia may each represent different features along a continuum of variable cerebellar involvement. Future natural history studies will be needed to determine whether cerebellar ataxia eventually develops in all patients. Nevertheless, the identification of a more limited phenotype in a sizeable number of patients suggests that the overall frequency of GAA-*FGF14* disease may even be higher than previously estimated.

While BVP has been suggested to be a recurrent feature of GAA-*FGF14* disease,^15^ our study is the first to provide an estimate of the frequency and temporal evolution of BVP in a large and phenotypically unselected cohort of patients with GAA-*FGF14* disease. Ancillary vestibular assessment performed in more than 95% of (GAA)_≥250_-*FGF14* patients documented BVP in 10% of them. BVP appears to be a late feature in GAA-*FGF14* disease compared to cerebellar ataxia, as it developed on average more than 10 years after disease onset. In addition to being significantly less frequent in (GAA)_≥250_-*FGF14* compared to (GAA)_<200_-*FGF14* DBN syndromes, BVP tended to remain relatively mild in GAA-*FGF14*-related DBN syndromes despite prolonged disease duration. The origin of the vestibular pathology in GAA-*FGF14* disease is yet to be established, although a central etiology may be more likely given the absence of significant involvement of the peripheral nervous system in this disease.

While previous studies of patients with SCA27B have shown rates of cerebellar atrophy ranging from 60 to 100%,^12,14^ only 13% of (GAA) -*FGF14* patients with DBN exhibited cerebellar atrophy (mainly of the vermis). This finding might stem from our cohort’s main inclusion criterion being DBN, not cerebellar ataxia, resulting in over half of the patients displaying no overt ataxia. Moreover, the median disease duration at time of brain MRI in our DBN cohort was relatively short (4.5 years). Mild cerebellar vermis atrophy can also be challenging to identify on routine MRI and may have been missed.

Leveraging in-depth phenotyping data, our study identified clinical features discriminating (GAA)_≥250_-*FGF14* from (GAA)_<200_-*FGF14* DBN syndromes. Of all clinical signs, impairment of the visual fixation suppression of the VOR was most strongly associated with and predictive of (GAA)_≥250_-*FGF14* DBN, also indicating a dysfunction of the cerebellar flocculus/paraflocculus.^28^ While multiple areas of the brain are involved in gaze stabilization during head movement, the cerebellar flocculus/paraflocculus is critical in modulating the VOR.^28^ Early dysfunction of these two structures of the vestibulocerebellum, which are also implicated in the pathophysiology of DBN,^3-6^ provides a unifying theory for the frequent and early co-occurrence of DBN, impaired VOR cancellation and other eye movement abnormalities, and vertiginous symptoms in GAA-*FGF14* disease.^29^

Functional impairment, as captured by the FARS functional disability stage and the need for walking aid, increased relatively slowly with disease duration in (GAA)_≥250_-*FGF14* patients, confirming previous reports from patients GAA-*FGF14* ataxia.^12,14^ The slow accrual of disability was evidenced by an increase of 0.10 FARS stage per year of disease and the need for mobility aids in 20% of patients. However, we observed inter-individual variability in the rate of disability progression among patients, which was at least in part accounted for by concurrent medical illnesses in some patients. These observations strengthen the previous hypothesis from patients with GAA-*FGF14* ataxia^14^ that co-morbid medical illnesses, which are common in elderly populations, must be taken into account when analyzing patient data and planning treatment trials in late-onset diseases like GAA-*FGF14* disease as they may alter their natural progression.

The pathogenic potential of (GAA)_200-249_ alleles has recently been suggested based on the observation of segregation of (GAA)_200-249_ alleles and (GAA)_≥250_ expanded alleles with the disease in an affected family with autosomal dominant late-onset slowly progressive cerebellar ataxia.^24^ Extending this preliminary evidence derived from segregation analysis, we now provide quantitative gene burden evidence for this notion by showing a significant enrichment of (GAA)_200-249_ alleles in patients with DBN compared to controls. In addition, as a clinical-phenotypic line of evidence, we observed that impairment of the visual fixation suppression of the VOR and treatment response to 4-AP, both strongly associated with (GAA)_≥250_-*FGF14* DBN syndromes, were significantly more common in (GAA)_200-249_-*FGF14* than (GAA)_<200_-*FGF14* patients (but not different from [GAA]_≥250_-*FGF14* patients). It might thus be conceivable that *FGF14* (GAA)_200-249_ alleles may cause a milder cerebellar phenotype more commonly manifesting with isolated DBN and cerebellar ocular motor signs. These findings highlight the need to conduct large-scale studies to re-evaluate the pathogenic threshold of GAA-*FGF14* disease.

Our study provides large scale evidence – including a first small set of placebo-controlled data – for the symptomatic benefit of 4-AP in patients with GAA-*FGF14* disease. Our findings showed that 4-AP treatment was associated with a clinician-reported treatment response in 80% and a patient-reported meaningful benefit in 59% of (GAA)_≥250_-*FGF14* and (GAA)_200-249_-*FGF14* patients. In comparison, such treatment responses were significantly lower in (GAA)_<200_-*FGF14* patients (31% and 11%, respectively). Although these data were collected without knowledge of the GAA-*FGF14* genotype but with open-label provision of treatment, the substantially lower response rate to 4-AP treatment in (GAA)_<200_-*FGF14* patients diminishes the likelihood that the observed effects can be largely attributed to placebo. Response to 4-AP in fact represented a strong predictor of (GAA)_≥250_-*FGF14* and (GAA)_200-249_-*FGF14* status in patients with idiopathic DBN (positive predictive value, 89%). The mechanisms by which 4-AP, a potassium channel blocker,^30^ ameliorates symptoms in GAA-*FGF14* disease are yet to be established although it may involve restoration of cerebellar Purkinje cell rhythmic firing property, as shown in other forms of hereditary ataxia,^30,31^ that is disrupted with the loss of *FGF14* function.^32,33^

Albeit still preliminary due to the small sample set, we present the first double-blind (and genotype-blind), placebo-controlled randomized treatment data of 4-AP in patients with GAA-*FGF14* disease.^16^ All four patients for whom DNA was available and who were found to carry an *FGF14* (GAA)_≥250_ repeat expansion showed an improvement of SPV of the DBN on 4-AP, but not placebo. This observation bolsters the existing open-label real-world evidence for the treatment efficacy of 4-AP in GAA-*FGF14* disease observed in our current cohort and in previous SCA27B cohorts.^14,18^ A larger randomized controlled trial is, nonetheless, warranted to confirm these findings. Our study highlights the potential of video-oculographic assessment of DBN as a responsive outcome measure for treatment. This approach could complement a promising battery of quantitative digital-motor outcomes,^18^ including sensor measures of gait and balance, in future trials of 4-AP for GAA-*FGF14* disease.

The results of our study need to be interpreted in light of some limitations. First, multi-centre studies with patient cohorts with DBN from different ethnic backgrounds are needed to replicate our findings, which were drawn from a single-centre patient cohort of overwhelmingly European background. Such studies are warranted to assess the frequency of *FGF14* GAA repeat expansions in other populations as profiling of the *FGF14* repeat locus in controls of diverse ancestries has shown that larger *FGF14* alleles are less common in non-European persons.^12,13^ Second, future prospective natural history studies in patients with DBN and other non-ataxia predominant phenotypes will be required to accurately track the phenotypic evolution of GAA-*FGF14* disease from its earliest stage and to establish whether a subset of patients never develops overt cerebellar ataxia. Third, our findings of potential pathogenicity of *FGF14* (GAA)_200-249_ alleles are largely preliminary and require validation through additional segregation studies, larger case-control series, and functional studies. Fourth, although we show a treatment effect of 4-AP in several outcomes, including clinician-reported and quantitative digital-motor outcomes, substantiating our real-world treatment response data, the benefits reported herein need validation in a larger randomized placebo-controlled trial.

In conclusion, we showed that *FGF14* GAA repeat expansions are a highly frequent monogenic cause of DBN syndromes in the European population. Our study suggests that GAA-*FGF14* disease may present along a continuum of variable cerebellar involvement, with some patients exhibiting isolated cerebellar floccular/parafloccular ocular motor signs without overt ataxia, and that *FGF14* (GAA)_200-249_ alleles may be associated with a disease phenotype. It also provides robust and large-scale evidence that the GAA-*FGF14* genotype status defines a subgroup of patients with DBN that appear to be highly responsive to 4-AP, further paving the way toward clinical trials.

## Data Availability

All data produced in the present study are available upon reasonable request to the authors.

## Author contributions

Design or conceptualization of the study: DP, CW, BB, MSt, MSy.

Acquisition of data: DP, FH, CW, MCD, AT, CA, MJD, AC, GDG, KMB, JC, DR, AMH, SZ, BB, MSt, MSy.

Analysis or interpretation of the data: DP, FH, CW, MCD, AT, CA, MJD, BB, MSt, MSy.

Drafting or revising the manuscript for intellectual content: DP, FH, CW, MCD, AT, CA, MJD, AC, GDG, KMB, JC, DR, AMH, SZ, BB, MSt, MSy.

## Declaration of interests

DP reports no disclosures.

FH reports no disclosures.

CW reports no disclosures.

MCD reports no disclosures.

AT reports no disclosures.

CA reports no disclosures.

MJD reports no disclosures.

AC reports no disclosures.

GDG reports no disclosures.

KMB reports no disclosures.

JC reports no disclosures.

DR has received grant/research support from Janssen and Lundbeck; he has served as a consultant or on advisory boards for AC Immune, Janssen, Roche and Rovi and he has served on speakers bureaus of Janssen and Pharmagenetix. He also received honoraria from Gerot Lannacher, Janssen and Pharmagenetix, and travel support from Angelini and Janssen, all unrelated to the present manuscript.

AMH reports no disculosures.

SZ has received consultancy honoraria from Neurogene, Aeglea BioTherapeutics, Applied Therapeutics, and is an unpaid officer of the TGP foundation, all unrelated to the present manuscript.

BB reports no disclosures.

MSt is Joint Chief Editor of the Journal of Neurology, Editor in Chief of Frontiers of Neuro-otology and Section Editor of F1000. He has received speakers honoraria from Abbott, Auris Medical, Biogen, Eisai, Gru[nenthal, GSK, Henning Pharma, Interacoustics, J&J, MSD, NeuroUpdate, Otometrics, Pierre-Fabre, TEVA, UCB, and Viatris. He receives support for clinical studies from Decibel, U.S.A., Cure within Reach, U.S.A. and Heel, Germany. He distributes M-glasses and Positional vertigo App. He acts as a consultant for Abbott, AurisMedical, Bulbitec, Heel, IntraBio, Sensorion and Vertify. He is an investor and share-holder of IntraBio. All are unrelated to the present manuscript.

MSy has received consultancy honoraria from Janssen, Ionis, Orphazyme, Servier, Reata, Biohaven, Zevra, Lilly, GenOrph, and AviadoBio, all unrelated to the present manuscript. MSy is planning a treatment trial of 4-AP in GAA-*FGF14* disease together with Solaxa Inc. as a sponsor, but has not received any type of honoraria or funding from Solaxa.

## Acknowledgments

We thank the patients and their families for participating in this study. We thank the Centre d’Expertise et de Services Génome Québec for assistance with Sanger sequencing. DP and GDG hold Fellowship awards from the Canadian Institutes of Health Research (CIHR).

